# Clinical validation of automated and multiple manual callosal angle measurement methods in idiopathic normal pressure hydrocephalus

**DOI:** 10.64898/2026.02.12.26346185

**Authors:** Woosung Seo, Shetha Jabur, Amir Rashid, Nils Holmstrand, Dag Nyholm, Johan Virhammar, David Fällmar

**Author notes:** Corresponding author: Woosung Seo MD.

## Abstract

**Introduction:** Idiopathic normal pressure hydrocephalus (iNPH) is a partially reversible neurological disorder in which imaging biomarkers support diagnosis and surgical decision-making. The callosal angle (CA) is one of the most robust radiological markers of iNPH and has also been associated with postoperative shunt outcome. However, several manual measurement variants exist and artificial intelligence (AI)-based tools now enable automatic CA measurement.

**Materials and Methods:** In total 71 patients (40 with confirmed iNPH and 31 controls) were included. Six predefined manual methods for measuring CA were applied to preoperative 3D T1-weighted MRI and evaluated for diagnostic performance and interobserver agreement. An AI-derived automatic CA (cMRI from Combinostics) was included as a seventh method and compared with the traditional manual method (perpendicular to the bicommissural plane and through the posterior commissure). Automatic measurements were additionally assessed in pre- and postoperative scans to evaluate robustness against shunt-related artifacts.

**Results:** All seven CA variants significantly differentiated iNPH patients from controls (p < 0.05). The traditional method showed the highest discriminative performance (AUC = 0.986, SE = 0.012), while alternative planes demonstrated slightly lower accuracy (AUC range = 0.957–0.978). Interobserver agreement for manual measurements was good to excellent (ICC = 0.687–0.977). Automatic CA measurements showed excellent correlation with the traditional method, preoperative ICC = 0.92; postoperative ICC = 0.96.

**Conclusion:** Although several CA positions perform comparably, the traditional method remains marginally superior and is best supported by the literature. Automated CA measurements closely match expert manual assessment in pre- and postoperative imaging, supporting clinical implementation.

## Introduction

Idiopathic normal pressure hydrocephalus (iNPH), also known as Hakim disease (1), is a potentially reversible neurological disorder characterized by the classic triad of gait disturbance, cognitive decline, and urinary incontinence, often accompanied by ventriculomegaly on neuroimaging in the absence of pronounced cortical atrophy (2). Early recognition is crucial, as timely surgical intervention with shunting of cerebrospinal fluid can lead to substantial functional improvement (3, 4). Neuroimaging plays a central role in both diagnosis and surgical decision-making (5). Among the numerous imaging markers that have been proposed for iNPH (6-10), the callosal angle (CA) has emerged as one of the most robust and reproducible markers of iNPH (7, 11, 12) and several studies have shown that the CA can, to some extent, predict positive surgical outcome (13, 14).

The use of CA in iNPH was introduced by Ishii et al. in 2008, and was defined as a measurement on coronal images perpendicular to the anterior–posterior bicommissural plane, at the level of the posterior commissure (6). This is described as the “standard” and “traditional” method throughout this paper. Narrowing of the CA reflects the vertical (upwards) expansion of the lateral ventricles, which is characteristic for iNPH. While the standard measuring technique is well established and validated in many publications, it requires precise multiplanar reconstructions and dedicated workflow steps that may not be easily deployed in all clinical settings, and less experienced users may struggle with the angulation of the coronal image. It is well established that small variations in measuring technique have a large influence on the result (8, 15).

Alternative measurement planes aligned with easily identifiable anatomical landmarks have been proposed to improve feasibility in routine practice (16-18). Whether these simplified methods maintain comparable diagnostic performance to the standard approach remains an open question, and interobserver variability across techniques is another consideration.

Recent advances in artificial intelligence (AI) offer new opportunities for standardizing and automating CA measurements. AI-based tools can detect anatomical landmarks, reconstruct optimal imaging planes, and compute the CA without manual input, thereby reducing both effort and interobserver variability, potentially improving workflow efficiency (19). If validated, such methods would facilitate rapid, consistent CA assessments across institutions, including those without specialized neuroradiology expertise. Hence it could be integrated into large-scale screenings or decision-support systems for iNPH.

Given these developments, rigorous clinical validation of both manual and automated CA measurements is essential before widespread implementation. Such validations should assess diagnostic accuracy in distinguishing iNPH, reproducibility between raters, agreement between automated and expert-derived measurements, and robustness in postoperative scans. This study aims to give a comprehensive comparison between different CA measurement techniques, including novel simplified planes and an AI-based automatic method, in order to provide evidence-based recommendations for their clinical use and to explore the potential of automation in improving the diagnostic workup and management of iNPH.

This study compares the discriminatory performance of six manual CA measurement techniques, including the established traditional method and several alternative methods including simplified approaches. A second objective was to validate an AI-based method for fully automatic CA quantification by assessing its agreement with expert manual measurement across pre- and postoperative MR imaging.

## Materials and Method

In the first step, 71 participants were included: 40 with confirmed iNPH who subsequently underwent shunt surgery, and 31 age-matched controls. Six manual methods to measure CA (hence named A–F) were evaluated on retrospectively collected preoperative 3D T1-weighted MRI scans. Because of the retrospective study design, there were some variances in imaging technique, but all images used in the study are MPRAGE sequences with 1 x 1 x 1 mm isotropic voxels, and with an image quality deemed sufficient for the intended purpose. The coronal planes and positions for manual measurements were predefined accordingly (also visualized in Figure 1).

**Figure 1.**
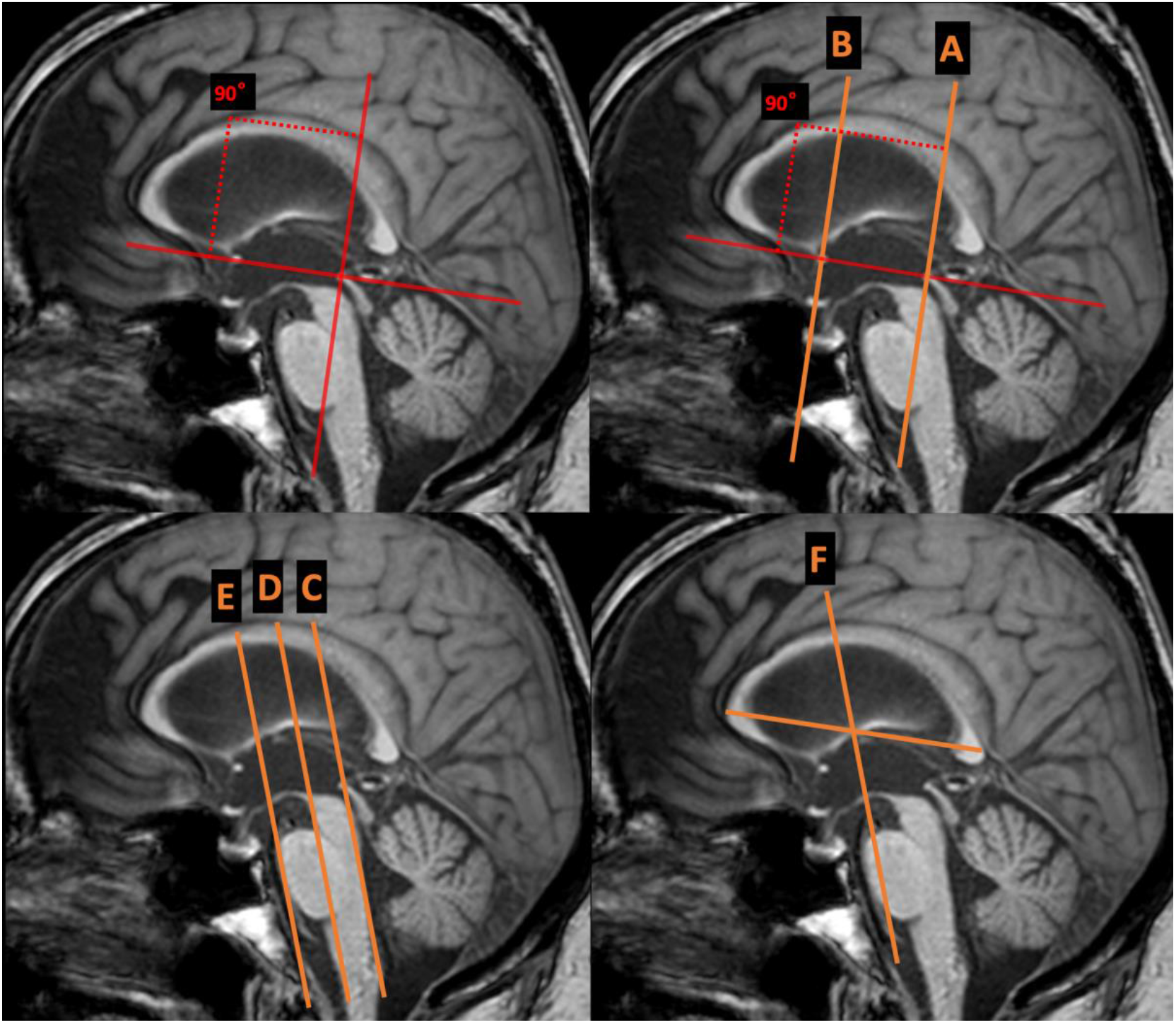
Different planes A-F shown on mid-sagittal images in which the callosal angle was subsequently measured manually on coronal MPRAGE images. The automatic measurement is intended to simulate the traditional position (A).

**Position A:** The traditional and established position (ad modum Ishii); in a coronal plane perpendicular to a line drawn through the centres of the anterior and posterior commissures (AC–PC), in a position through the centre of the posterior commissure (6).

**Position B:** In a coronal plane perpendicular to the AC–PC plane (as A), but in a position through the centre of the anterior commissure, as proposed by Mantovani et al (17).

**Position C:** Coronal plane along the dorsal pons (conventional coronal plane, according to many centres), and also in the position of dorsal pons.

**Position D:** Coronal plane along the dorsal pons (as C), but in the position of central pons. **Position E:** Coronal plane along the dorsal pons (as C+D), but in the position of its anterior border.

**Position F:** The “simplified” CA, according to Cagnin et al.; in a coronal plane along the dorsal pons, in a position half-way between the anterior and posterior borders of the corpus callosum (16).

A consensus protocol for manual assessments was developed and piloted before data collection to ensure consistency. A subset of measurements was repeated by an experienced, board-certified neuroradiologist (DF), to assess inter-rater agreement.

As a second step, fully automated measurements of the CA were acquired from the same images, using the commercially available cNeuro cMRI software, version 2.3.4 (Combinostics OY, Tampere, Finland). These were handled as a seventh variant of CA measurement method (named G), but the software is designed to replicate the traditional method (here named position A). Sample images are shown in Figure 2. The cMRI software was installed on-premise and integrated in the research-PACS environment.

**Figure 2.**
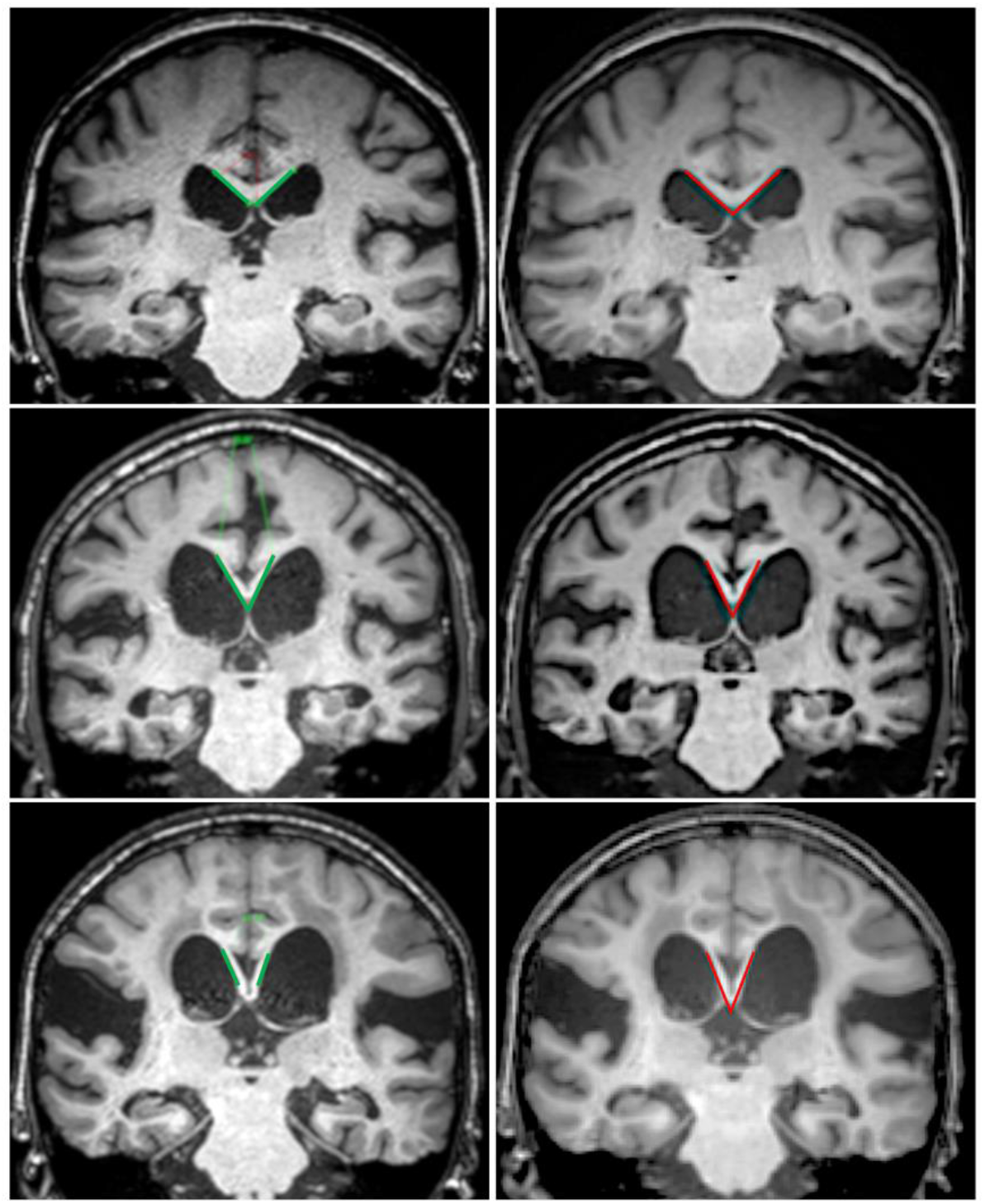
Figure above shows three randomly chosen comparisons between manual and automatic measurements of the callosal angle. The left column shows manual measurements in PACS, using a power viewer mode with free MPR positioning and the traditional method of Ishii et al. (position A in this study). The right column shows automatic output from the cMRI software. The upper row shows a healthy control, and the two bottom rows show iNPH patients with pathologically sharp angles. The congruency between methods is dependent both on the placement of the angulation lines, and the positioning of the underlying coronal planes.

**Figure 3.**
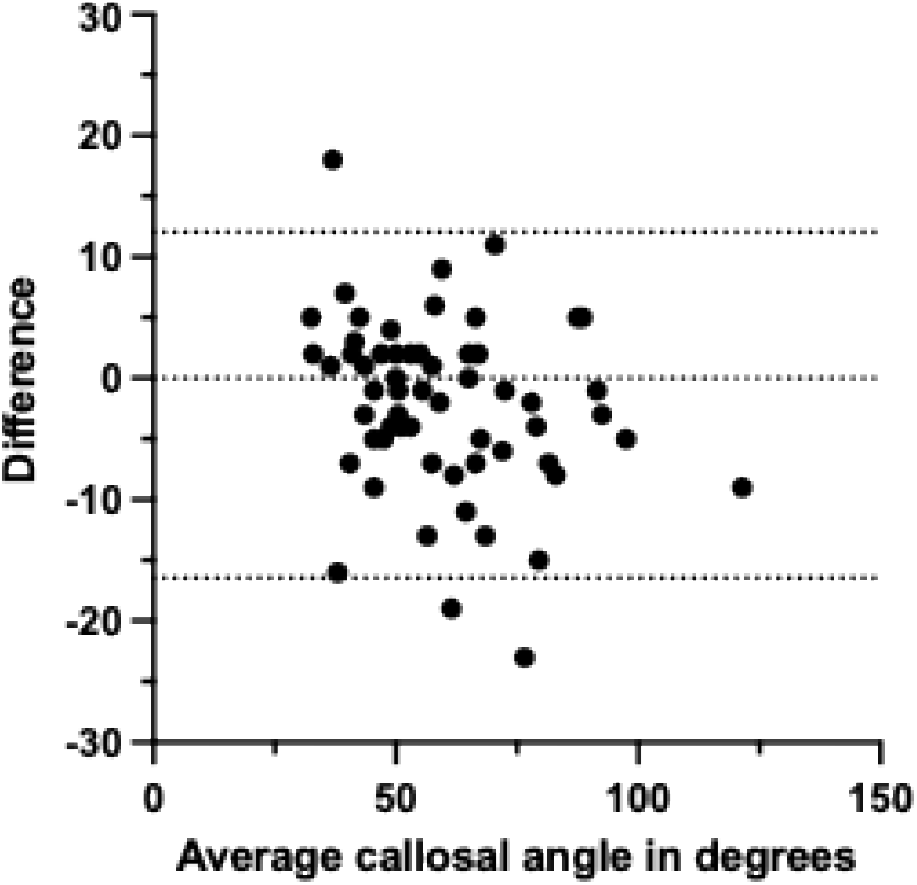
Bland-Altman analysis of automated and manual callosal angle measurements. The plot shows the difference between automated and traditional manual callosal angle measurements plotted against their mean in preoperative MRI. Most measurements fall within the 95% limits of agreement (dotted lines), indicating excellent concordance between the methods.

For the third step of the study, two additional cohorts were added for a larger validation of the automatic measurements from the cMRI software. The automatic method was compared to the results from position A in the first step. Preoperative scans from 39 iNPH patients were included from other ongoing research projects. Further, a subset of 19 iNPH patients were scanned postoperatively as a follow-up after ventriculoperitoneal shunt insertion (with programmable shunt valves). These were included in a separate subgroup to validate the robustness of the automatic measurements in the presence of image artifacts from the shunt valves. These two additional cohorts were hence analysed using only the automatic and traditional (position A) CA measurements.

### Statistical Analysis

Statistical analyses were performed to evaluate discriminatory performance across the seven measurement methods. Discrimination between iNPH patients and controls were assessed using receiver operating characteristic (ROC) analysis, with calculation of the area under the curve (AUC) and corresponding standard error. Inter-rater reliability for the manual CA measurements was quantified using a two-way mixed-effects intraclass correlation coefficient. Agreement between automated CA measurements and the traditional manual method (Position A) was further assessed using ICC for both preoperative and postoperative MRI datasets.

A Bland–Altman analysis was performed exclusively for the preoperative dataset to evaluate systematic bias and 95% limits of agreement between the two methods. Ninety-five percent confidence intervals (95% CI) were calculated for ICC and Bland–Altman estimates. Statistical significance was defined as a two-sided p < 0.05.

Statistical analyses and figure generation were performed using Microsoft Excel and GraphPad Prism (version 10.6.1; GraphPad Software, San Diego, CA).

## Results

All seven CA methods significantly discriminated patients from controls (p < 0.05). As shown in Table 1, the traditional manual method (perpendicular to AC-PC plane in PC position, position A) showed the highest performance and the smallest standard error (AUC = 0.986 and SE = 0.012). Alternative (including “simplified”) methods had similar but slightly lower performance (range of AUC 0.957–0.978). Inter-rater agreements were good to excellent (ICC range 0.687–0.984), with the highest coefficient seen for the traditional method. The cMRI-derived automatic method showed a discriminatory performance comparable to the manual technique (AUC = 0.958) and excellent agreement to the traditional CA method (ICC = 0.92).

**Table 1.** Diagnostic performance and inter-rater agreement of callosal angle variants.

| *Callosal angle variants* | *Area under the curve (AUC)* | *Standard Error (SE)* | *Intraclass correlation coefficient (ICC)* |
| --- | --- | --- | --- |
| *Position A* | *0.986* | *0.012* | *0.984* |
| *Position B* | *0.964* | *0.024* | *0.892* |
| *Position C* | *0.957* | *0.026* | *0.918* |
| *Position D* | *0.972* | *0.021* | *0.960* |
| *Position E* | *0.963* | *0.026* | *0.687* |
| *Position F* | *0.978* | *0.019* | *0.977* |
| *Position G (cMRI)* | *0.958* | *0.022* | - |

**Table 1.** Area under the curve (AUC) from receiver operating characteristic analysis shows discriminatory power between idiopathic normal pressure hydrocephalus (iNPH) and controls. Positions A–F represent manual callosal angle measurements, while position G denotes automatic measurements (cMRI). Intraclass correlation coefficient (ICC) reflects inter-rater agreement for manual measurements.

After adding the cohort with an additional 39 preoperative patients for further validations of automatic measurements to the traditional method (position A), the combined comparisons yielded a result with ICC value of 0.92 for preoperative cases (95% CI: 0.88 – 0.95). For the subset of postoperative patients, automatic CA continued to show excellent agreement with ICC 0.96 (95% CI: 0.89 – 0.98) with no signs of artifact-derived decrease in CA delineation.

## Discussion

One key finding in this study is that several CA measurement techniques, including “simplified” planes, provide high performance in distinguishing iNPH patients from controls - with the traditional method of Ishii et al. remaining slightly superior. A second key finding is that a strong correlation between the AI-derived automatic CA measurements and the manual gold standard, in both pre- and postoperative scans, underscores the potential of automated tools to enhance reproducibility and efficiency in clinical workflows.

Based on the subjective experience of performing the manual measurements, none of the alternative methods was perceived as being clearly “simpler” than the well-defined standard method (A). In fact, the proposed “simplified CA” of Cagnin et al., here named position F, was considered more difficult and time-consuming since defining half the length of the corpus callosum is a necessary step to determine the correct position (16).

This study has several limitations. The study cohort was derived from a single centre which may limit generalizability. Furthermore, although the AI tool performed well overall, occasional discrepancies were observed in cases with atypical ventricular morphology or severe white matter changes, suggesting that algorithmic refinements and larger-scale validation may be needed. Also, an optimal study of clinical usefulness should compare patients to other patients with mimicking diseases rather than (or in addition to) healthy controls. For this methodological validation study with a straight-forward measuring approach, the drawback from only comparing to healthy controls was considered acceptable. Future research should assess the clinical impact of automated CA measurement in diverse imaging environments (including real-life settings using control groups with relevant similar diseases), explore its integration with other quantitative biomarkers, and evaluate its role in longitudinal monitoring and surgical outcome prediction in iNPH.

## Conclusion

This study demonstrates that although several callosal angle (CA) measurement techniques yield high accuracy for distinguishing iNPH from controls, the traditional method remains marginally superior and is best supported by the current literature. Accordingly, adopting alternative CA planes should be justified by clear clinical or workflow benefits rather than convenience alone. Further, automatic measurements using a commercially available tool showed excellent agreement with the traditional method, even in postoperative scans despite shunt-related artifacts.

Collectively, these findings support the clinical usefulness of automated CA measurements. By minimizing interobserver variability, supervised automated assessments can strengthen the routine diagnostic workup of iNPH, especially in centres without specialised expertise.

## Data Availability

All data produced in the present study are available upon reasonable request to the authors.

## Notes

### Competing Interest Statement

The authors have declared no competing interest.

### Funding Statement

D.Fallmar was supported by the Swedish Brain Foundation (Grant No. PS2021-0026) and the Swedish Society for Medical Research (SSMF, Grant No. PD21-0136). J.Virhammar was funded by the Swedish Society for Medical Research (SG-22-0192-H-01).

### Author Declarations

The ethical approval is covered in the EPM approvals Dnr 2010/161/1-2 and Dnr 2015/174/1-4, with addendums 2021 and -22. The study was approved by the Swedish ethical approval authorities.

